# Modelling the epidemic growth of preprints on COVID-19 and SARS-CoV-2

**DOI:** 10.1101/2020.09.08.20190470

**Authors:** Giovani L. Vasconcelos, Luan P. Cordeiro, Gerson C. Duarte-Filho, Arthur A. Brum

## Abstract

The response of the scientific community to the global health emergency caused by the COVID-19 pandemic has produced an unprecedented number of manuscripts in a short period of time, the vast majority of which have been shared in the form of preprints posted on online preprint repositories before peer review. This surge in preprint publications has in itself attracted considerable attention, although mostly in the bibliometrics literature. In the present study we apply a mathematical growth model, known as the generalized Richards model, to describe the time evolution of the cumulative number of COVID-19 related preprints. This mathematical approach allows us to infer several important aspects concerning the underlying growth dynamics, such as its current stage and its possible evolution in the near future. We also analyze the rank-frequency distribution of preprints servers, ordered by the number of COVID-19 preprints they host, and find that it follows a power law in the low rank (high frequency) region, with the high rank (low frequency) tail being better described by a *q*-exponential function. The Zipf-like law in the high frequency regime indicates the presence of a cumulative advantage effect, whereby servers that already have more preprints receive more submissions.

## 1 INTRODUCTION

As of this writing, more than 110 millions of cases of infection by the novel coronavirus (SARS-CoV-2) have been identified worldwide and nearly 2.5 millions of deaths have been attributed to the ensuing disease (COVID-19) [1, 2]. In its wake, the COVID-19 pandemic has seemingly touched, in one way or another, every aspect of our daily lives. Not surprisingly, given the health risks represented by the SARS-CoV-2 virus and the many scientific challenges involved, the COVID-19 pandemic has had a huge impact on the scientific community. On the downside, social isolation and lockdown measures, for example, have caused disruption (if only temporary) of research projects that were being undertaken before the onset of the crisis and made it nearly impossible for researchers to collaborate and discuss their work in person. But on the up side, the scientific community responded quickly to the challenges posed by the unprecedented crisis by producing an unprecedented number of scholarly works in a short period of time.

A considerable proportion of this scientific output has been disseminated in the form of preprints posted before peer review on open and publicly available online platforms. Despite some concerns regarding the quality of preprints [3, 4, 5, 6, 7], in comparison to peer-reviewed articles, there seems to be a growing consensus that the benefits of the rapid sharing of information allowed by preprints “far outweigh the disadvantages” [4]; see also Ref. [8] for a recent discussion (before the onset of COVID-19) of the advantages of and problems with preprints in medicine and the biological sciences—two areas that were slower in adopting preprint practices. Many studies about both peer-reviewed papers and unrefereed preprints related to COVID-19 research have recently been conducted, but mostly with emphasis on the bibliometrics aspects of the topic; see, e.g., Refs. [9, 10, 11, 12, 13, 14, 15] and references therein.

In the present paper we take a different approach from these previous studies. Here we seek to understand the growth dynamics underlying the rapid surge of preprints related to COVID-19 research. To this end, we employ a generalized logistic growth model to describe the time evolution of the cumulative number of preprints deposited on preprint repositories. Since the pioneering work by Verhulst on the standard logistic model [16], phenomenological growth models have been successfully applied to many growth processes, ranging from human [16] and animal [17] population dynamics to epidemics [18], including the COVID-19 epidemic itself [19, 20]. It is thus natural to expect that growth models should also be applicable to this “epidemic in an epidemic,” as the rapid surge of preprints on COVID-19 has been referred to in Ref. [13]. We shall also use the shorter term “scidemic” for this phenomenon.

More specifically, here we apply the generalized Richards model (GRM) to describe the cumulative curve of COVID-19 preprints. We show that the GRM does give a very good fit to the empirical curve. Furthermore, the model allows us to extract relevant information about the social dynamics driving this quick growth of preprints. We show, for instance, that the early growth in the number of COVID-19 preprints follows a subexponential regime—rather than an exponential behavior, as was initially thought [9, 15, 21]. The model also predicts that after reaching a maximum growth rate (i.e., zero acceleration) in late May, 2020, the growth dynamics entered a decelerated phase that reached its maximum deceleration by late July, 2020, after which it moved into a regime of decreasing deceleration. We remark, parenthetically, that in October, 2020, there was a small increase in the rate of submissions of COVID-19 preprints, which might be an indication of a possible “second wave” in the scidemic, mimicking perhaps what happened in the actual COVID-19 epidemic [22]. As it is necessary to wait for more data to accrue to test this hypothesis, possible second wave effects will not be pursued here.

We also analyze the distribution of the COVID-19 preprints among the many available servers. We find that the corresponding rank-frequency distribution follows a power law (at least for the first few largest servers), similar to the Zipf law found in many preferential attachment processes, such as the frequency of words in a text [23], the size distribution of cities [24], the wealth distribution of individuals [24], the number of scientific citations [25], the number of joint publications [26], and the distribution of nodes in social networks [27], among others. We thus argue that the distribution of manuscripts among repositories also seems to follow a similar preferential attachment dynamics, whereby servers that already have more preprints tend to get more submissions. At the high rank (low frequency) tail of the distribution, a *q*-exponential function appears to give a better description to the data; which is an interesting result in view of the fact *q*-exponential distributions have been found in many complex systems (see, e.g., [28] and references therein), including in the context of scientific citations [29].

Our results show that the rapid multiplication of preprints on COVID-19 is a direct manifestation of a complex social dynamics that stems from a sort of “contagion process,” whereby existing preprints tend to spur more preprints, and so on. Modeling the underlying “microscopic” dynamics of this process is of course an interesting but daunting task, because the complex interaction between the agents, namely the scientific authors, is mediated by the appearance of the preprints themselves, rather than by the direct contact among authors. The alternative approach we adopt here, in terms of a mathematical growth model, has the advantage that, by avoiding the description of social mechanisms that may be difficult to identify, it allows us to make predictions about the resulting dynamics in a quantified manner. Our study thus helps to shed new light on the complex growth dynamics of COVID-19 preprints and related problems.

## 2 MATERIALS AND METHODS

### 2.1 Data Source

The data used in this study were obtained from the GitHub site maintained by Fraser and Kramer [30]. As explained in their site, preprints are considered to be related to COVID-19 on the basis of keywords matches in their titles or abstracts, according to the following search string: coronavirus OR covid-19 OR sars-cov OR ncov-2019 OR 2019-ncov OR hcov-19 OR sars-2. Their dataset is not meant to be exhaustive, but it collates information from an impressive list of 38 preprint servers [30]. As explained in [30], “only the earliest posted version” of a preprint is included in the dataset. Furthermore, in cases where a preprint is deposited in multiple repositories, “all preprint records are included.”

The first preprint included in their collection was posted on bioRxiv on January 15, 2020 [31]. The dataset used in the present study was updated up until September 30, 2020, and contains a total number of 28,757 preprints, distributed among 38 preprint servers, as already mentioned. We considered data only up to this date, to avoid possible second wave effects, as the model is designed to describe only one-wave growth profiles (i.e., sigmoidal curves). For the list of the twenty-one largest servers (as per the number of COVID-19 preprints), see Sec. 3. A complete list of the preprint repositories in the dataset can be found in [30].

It is important to recall that manuscripts deposited in preprint repositories are not certified by peer review. Nonetheless, they are subjected to a screening process by the respective servers administrators to ensure a minimum quality control. Preprint servers differ somewhat in their screening procedures, but they all seek to enforce guidelines against “poor science,” especially in the context of an emergency health crisis such as the COVID-19 pandemic [6]. Many manuscripts are first posted on a preprint server before (or simultaneously as) being submitted to a regular peer-reviewed journal. Other submissions are supposedly intended only as preprints and are never submitted for formal publication [32]. Here we make no distinction between these two cases, as this would require a much more extended research, which would be made even more challenging by the often long delay between submission of a manuscript to a peer-reviewed journal and its final publication. There are also articles that appear only in peer-reviewed journals and are not posted previously as preprints—hence they are not detected by searches in preprint repositories—, but these are admittedly becoming less frequent today [6]. As already noted, duplicated preprints (i.e., deposited in more than one server) are all included in the database, and so they are counted more than once, but arguably they appear in small numbers so as not to significantly affect the total statistics. Notwithstanding these small imperfections in the database, the growth dynamics of research preprints related to COVID-19 is a very good proxy for the response by scientific community to the crisis posed by the novel coronavirus.

### 2.2 Mathematical Growth Model

We model the growth dynamics by means of the generalized Richards growth model (GRM), which is defined by the following ordinary differential equation (ODE):

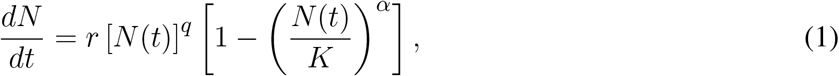

where *N* (*t*) is the growing quantity at time *t*; *r* is the growth rate at the early growth stage; *q* controls the initial growth regime and allows to interpolate from linear growth (*q* = 0) to sub-exponential growth (*q <* 1) to purely exponential growth (*q* = 1); *α* is the asymmetry parameter that controls the asymmetry of the growth profile with respect to the symmetric S-shaped curve of the logistic model, which is recovered for *q* = *α* = 1; and *K* represents the total quantity at the end of the growth process (i.e., *N* (*t*) = *K* for *t → ∞*). Equation (1) must be supplemented with the initial condition *N* (0) = *N*_0_, for some given value of *N*_0_.

Here we shall apply the GRM to the growing number of COVID-19 related preprints, so that *N* (*t*) will represent the cumulative number of preprints in our dataset up to the time *t*, where *t* is measured in days since the first preprint (hence *N*_0_ = 1). In adjusting the GRM to the empirical data we need to determine four free parameters, namely (*r, q, α, K*); the numerical fit is made easier by the existence of an analytical solution for the GRM, as discussed next.

Equation (1) admits an exact solution in implicit form given by [20]

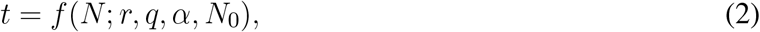

where

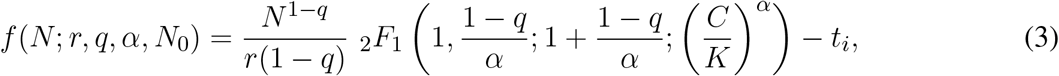

with _2_*F*_1_(*a, b*; *c*; *x*) denoting the Gauss hypergeometric function [33] and

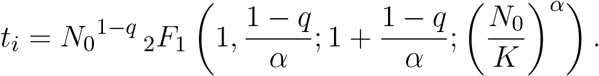

The fact that the solution of the GRM is given implicitly as *t*(*N*), rather than as an explicit function *N* (*t*), does not represent any hurdle to its practical use. Indeed, the above solution can be directly applied for curve-fitting purposes by viewing the empirical data in the same ‘implicit’ form, namely *t*_*k*_ as a function of *N*_*k*_, where *N*_*k*_ are the data points at times *t*_*k*_. The availability of an exact solution also has the advantage that it allows us to compute explicitly the location of certain key characteristic points of the growth profile, as indicated below.

For example, the inflection point *t*_*c*_ of the curve *N* (*t*), where 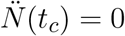, is given by [20]

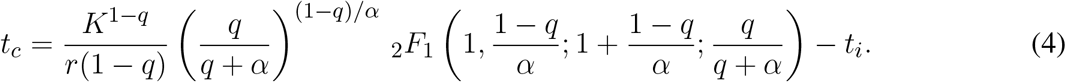

Knowledge of the inflection point *t*_*c*_ is important because it divides the growth process into two main phases according to its acceleration, as follows: i) an accelerating phase, for *t < t*_*c*_, when 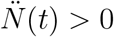 and ii) a decelerating phase, for *t* > *t*_*c*_, during which 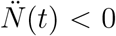. Each of these two main phases can be further divided into two subphases, according to whether the corresponding acceleration/deceleration is increasing or decreasing. More specifically, rec.a..lling that the rate of acceleration is known as the jerk, let us denote the points of zero jerk by 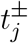, with 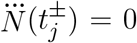, where 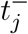 and 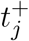 correspond to the points of maximum acceleration and maximum deceleration, respectively, so that 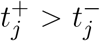. After some tedious algebra one finds that 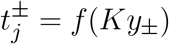, where *f* (*x*) is as given in (3) and

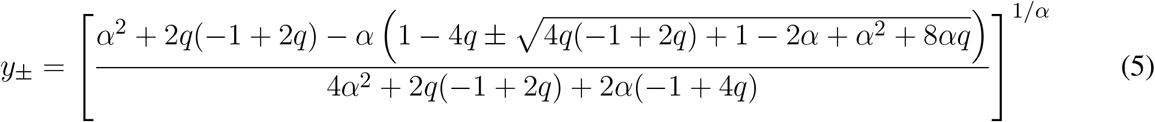

Now, comparing the time, *t*_*f*_, of the last empirical datapoint (corresponding to the ‘current time’) with the characteristic points 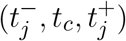 of the theoretical curve allows us to classify the current stage of the growth process. More specifically, we can define four growth stages, as follows: i) increasing acceleration, if 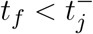 ii) decreasing acceleration, if 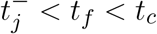 iii) increasing deceleration, if 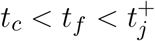 and iv) decreasing deceleration, if 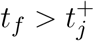. Having such a finer ‘diagnosis’ of the growth process is useful not only because it provides valuable information about the stage of the underlying dynamics at a given time, but also because it allows us to make predictions about its likely evolution within the near future. For instance, depending on how close the last time *t*_*f*_ is in comparison to the nearest phase-separation point, we may have an idea of how recently the growth curve has entered its current stage or how soon it may transition to the next one (if it is not in the final stage).

In particular, we note that in the first subphase, i.e., for 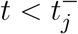, the GRM predicts a polynomial growth of the form [20]

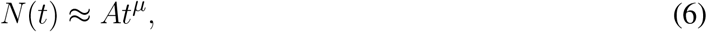

where *A* = [*r*(1 −*q*)]^1*/*(1−*q*)^ and *µ* = 1*/*(1 −*q*), for *q <* 1. In contradistinction, early exponential growth is obtained only for *q* = 1, in which case one has *N* (*t*) *≈ N*_0_ exp(*rt*). Similarly, in the late-time dynamics, i.e., for 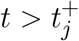, the GRM predicts an exponential rise to the plateau of the form [20]: *N* (*t*)−*K* ∝ exp(−*γt*), where *γ* = *rα/K*^1−*q*^.

### 2.3 Rank-Frequency Distribution

To analyze the size distribution of the preprint repositories, we first order the repositories according to the number of COVID-19 preprints they hold, where *n* = 1 is attributed to the repository with the highest number of preprints, *n* = 2 for the second largest repository, and so on. The relative frequency of preprints in the *n*-th repository will be denoted by *P* (*n*), where *P* (*n*) = *N*_*P*_ (*n*)*/Σ*_*j*_ *N*_*P*_ (*j*), with *N*_*P*_ (*j*) denoting the number of preprints in the *j*-th largest repository.

Preprint servers operate largely under similar principles. They aim to provide a publicly and freely accessible platform for rapid dissemination and sharing of scientific manuscripts that were not yet certified by peer review. There are preprint repositories that specialize in certain areas, such as: arXiv for physics and mathematics; bioRxiv and medRxiv for biomedical sciences; SSRN for the social sciences; and RePEc for economics research. There are also multidisciplinary preprint platforms, such as Research Square, OSF Preprints, and others. As already mentioned, there are 38 preprint servers in our dataset, with 21 of them hosting more than 40 COVID-19 manuscripts.

Authors thus have a substantial array of preprint servers to choose from when submitting a manuscript on COVID-19 related research. This is in quite contrast to non-COVID-19 preprints, where authors often prefer to use servers that specialize in their disciplines. It is therefore interesting to investigate how authors decide to which server to submit their COVID-19 preprints. For instance, preprint repositories may differ somewhat in their screening procedures and other policy requirements [3, 6], but it is fair to argue that these eventual differences do not represent a significant decision factor. In other words, the ‘cost’ of submission (say, in terms of time and extra work involved in the submission process) and the eventual ‘risk’ of rejection (if the manuscript is deemed not appropriate for the chosen server) are roughly the same among the different platforms. It is therefore reasonable to expect that authors will preferentially seek those platforms that may provide greater visibility for their work.

A reasonable decision strategy in such situation is of course to favor those repositories that already have a substantial number of preprints. In the context of the COVID-19 emergency, this strategy makes particular sense for authors whose principal areas of expertise are not directly related to, say, epidemiology, infectious diseases, virology, etc. This selection dynamics naturally leads to the so-called cumulative advantage [34] or preferential attachment processes [27], whereby those repositories that already have more preprints receive more. Preferential attachment processes usually lead to rank-frequency distributions, *P* (*n*), that exhibit power-law decay or the so-called Zipf law [23, 24, 34, 35]:

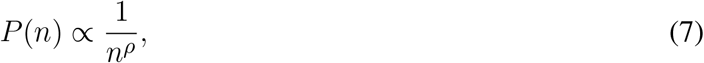

for *n* ≥ 1 and *ρ* > 0.

Zipf-like laws are particularly ubiquitous in linguistics and social processes, such as the frequency of words in a text [23], the size distribution of cities [24], the wealth distribution of individuals [24], the number of scientific citations [29], the distribution of degree of nodes in social networks [26, 36, 37], and the distributions of the tags in collaborative tagging networks [38], among others. The exponential function has also been applied to rank-size distributions, such as for urban and rural settlements; see, e.g., Ref. [39] and references therein. Below we discuss a general formalism that yields both power-law and exponential rank-size distributions as particular cases.

We formulate the rank-size distribution as a growth model described by the following ODE:

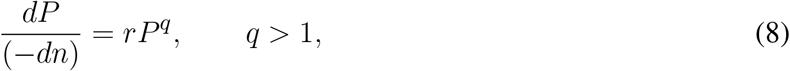

where *r* is positive constant. As written in the more unusual form above (with the negative sign on the left hand side), Eq. (8) can be viewed as describing the growth of the relative size, *P* (*n*), of the class of rank *n*, as *n* decreases (i.e., for *dn <* 0). Seeing the emergence of the rank-size distribution as a growth dynamics is not only in keeping with the general approach of the present work, but it also provides an effective model to derive the distribution *P* (*n*). (This innovative approach will be explored in a broader context in a forthcoming publication.) In this perspective, the exponent *q* in (8) can be interpreted as a measure of the “degree” of preferential attachment, in the sense that *q* = 1 implies a standard exponential decay for increasing *n* (or, alternatively, an exponential growth towards smaller *n*); whereas *q* > 1 corresponds to a slower than exponential decay for increasing *n*. Hence the larger the value of *q*, the greater the deviation from exponential behavior and thus the larger the degree of preferential attachment. (Strictly speaking, *n* is an integer variable, but if the number of ranks is large the continuous limit adopted in (8) is a valid approximation.)

The general solution of (8) is

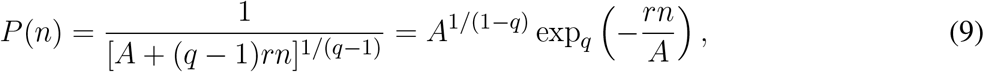

where exp_*q*_ (*x*) = [1 + (1 −*q*)*x*]^1*/*(1−*q*)^ is the *q*-exponential function [28, 29] and *A* is a constant of integration that can be fixed with a suitable initial condition. Here there are two cases of interest: i) *P* (0) *→ ∞*, which yields *A* = 0, in which case the distribution *P* (*n*) reduces to a pure power law:

*P* (*n*) = *Cn*^−*ρ*^, where *C* = 1*/*[(*q —*1)*r*]^1*/*(*q*−1)^ and *ρ* = 1*/*(*q* −1); and ii) *P* (0) *< ∞*, which implies *A* > 0, thus leading to power-law decay only for large *n*, that is, *P* (*n*) ∝ *n*^−*ρ*^, for *n »* 1, whereas *P* (*n*) approaches a constant for small *n*: *P* (*n*) ∝*n*^0^, for *n →*0. Clearly, the case *q* = 1 yields an exponential rank-size distribution: *P* (*n*) = *A* exp (−*rn*). One may also accommodate the possibility of two regimes in the ranked data [40] by allowing the small and large *n* regimes to be described by different exponents, namely, *q*_1_ (with *A*_1_ = 0) and *q*_2_ (with *A*_2_ > 0), for the low and high rank regions, respectively. In Sec. 3, we shall investigate the frequency distribution of COVID-19 preprints by repositories in light of the preceding discussion.

### 2.4 Statistical Fits

To perform the statistical fit for the GRM, we employed the Levenberg-Marquardt algorithm to solve the non-linear least square optimization problem, as implemented in the *lmfit* package for Python [41], which provides the parameter estimates and their respective errors. Here we have set *N* (0) = *N*_0_ = 1, so that according to (2) we are left with four parameters, namely (*r, q, α, K*), to determine numerically. In the case of the rank-frequency distribution, we fitted the selected data with power-law, exponential, and *q*-exponential functions, and also used the *lmfit* package to determine the free parameters in each case. The computer codes for the statistical fits were written in the Python language, and the plots were produced with the data visualisation library *Matplotlib*.

## 3 RESULTS

In Fig. 1 we show the cumulative number (red circles) of preprints on Covid-19 in our dataset, which we recall covers from January 15, 2020, to September 30, 2020, together with the GRM best fit (black solid curve). One sees from this figure that the theoretical curve describes very well the empirical data. Also shown in Fig. 1 are the point of maximum acceleration (orange vertical line), the inflection point (yellow vertical line), and the point of maximum deceleration (green vertical line), as obtained from the theoretical fit. The legend box in the figure shows the parameter estimates from the best fit, along with their respective errors.

**Figure 1.**
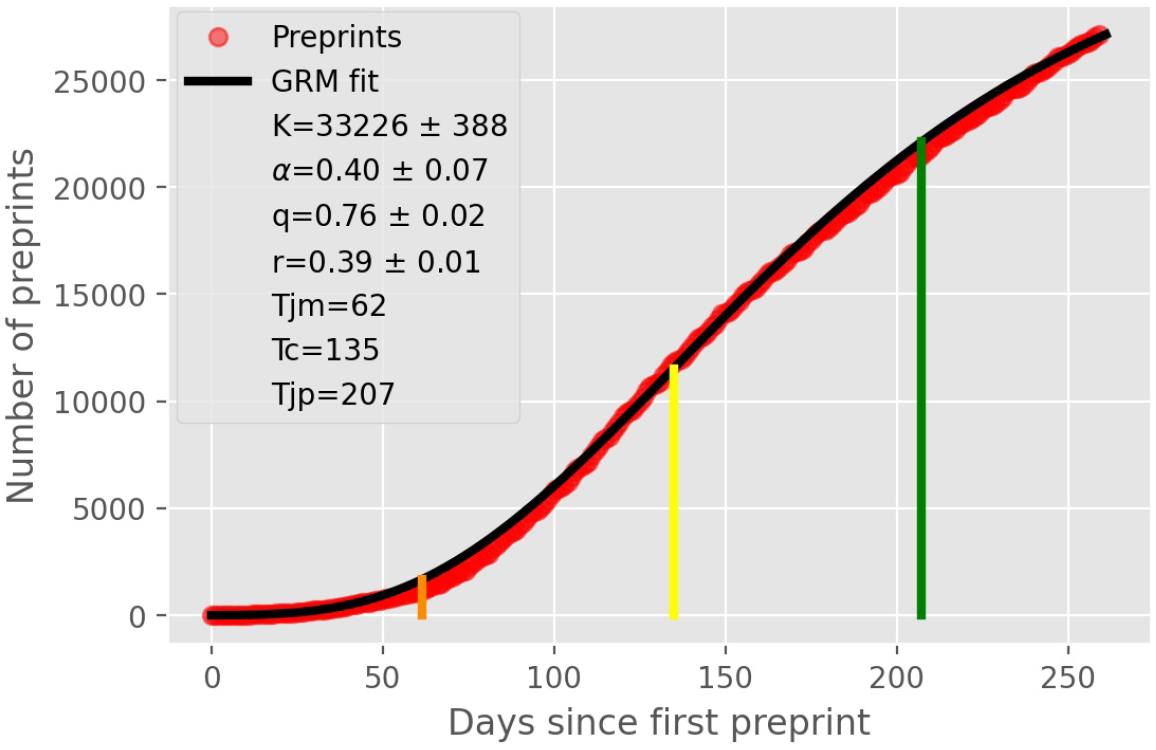
Cumulative number (red circles) of COVID-19 related preprints deposited on online preprint repositories up to September, 30, 2020. The solid black curve is the fit to the empirical data by the generalized Richards model, with the parameters shown in the legend box. The vertical lines indicate the location of some key characteristic points of the theoretical curve, as follows: i) point of maximum acceleration 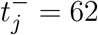; ii) inflection point *t*_*c*_ = 135 (yellow line); and iii) point of maximum deceleration 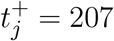.

In Fig. 2 we show the daily number of preprints (green curve) and the theoretical daily curve, which corresponds to the time derivative of the model cumulative curve shown in Fig. 1. Although the daily curve is quite noisy, as expected for a random-like process such as preprint submissions, we clearly see that the theoretical curve captures rather well the general trend of the daily data. In particular, the theoretical estimate for the “peak” of the daily curve (corresponding to the inflection point *t*_*c*_ of the cumulative curve) matches quite well the location of the region of largest values in the empirical curve.

**Figure 2.**
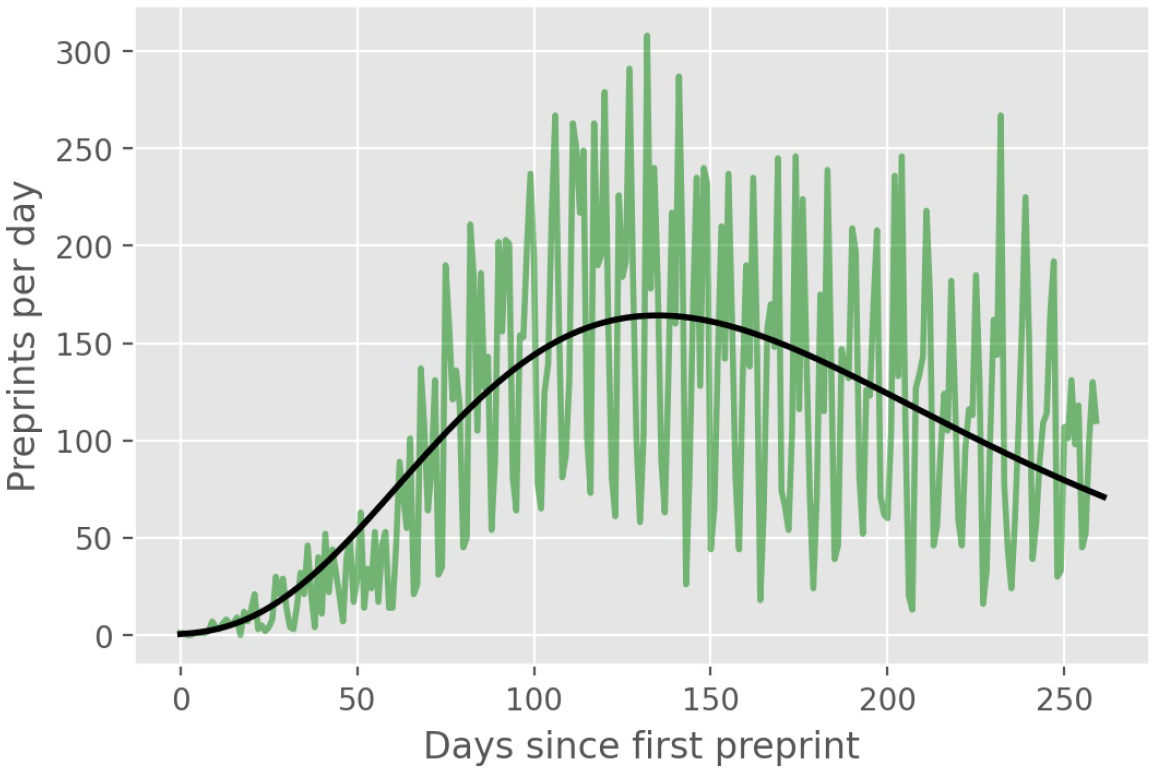
Daily number (green line) of COVID-19 preprints posted on online preprint repositories up to September, 30, 2020. The solid black line is the derivative of the theoretical cumulative curve shown in Fig. 1.

In the main plot of Fig. 3, we show the rank-frequency distribution for the preprint repositories that have at least 40 preprints; there are twenty-one of them, which account for the near totality (99.2%) of all COVID-19 related preprints. We note furthermore that the first ranked server (medRxiv) alone contributes with over one quarter (25.1%) of all submissions, which is twice the frequency of the second ranked repository (SSRN, with 12.6%). In the inset of Fig. 3, we show the complete rank-size distribution in log-log scale, together with the respective fits by an exponential function (red dotted line) and a *q*-exponential with *q* = 1.05 (black dashed line). One sees from the figure that the exponential function does not seem to provide a good description of the data at the high rank (low frequency) tail, in which case the *q*-exponential follows more closely the data. In the low rank (high frequency) end, say, for *n* ≤ 10, there is no distinction between the exponential and the *q*-exponential; but in this region the data points are slightly better described by a power law, as indicated by the green straight line, which is a power-law fit to the first seven data points. Taken together, these seven largest preprint servers (as per the number of preprints on COVID-19) account for nearly 80% of all COVID-19 preprints. Moreover, one sees in Fig. 3 that there is a noticeable ‘gap’ in size between the seventh (ResearchGate) and the eighth (JMIR) largest repositories, which suggests that there is indeed a change in dynamics at this point. These evidences appear to indicate that the seven largest preprint servers dominate (at least, up to the time of the present analysis) the ‘preferential attachment’ submission process. Evidence of two regimes in the ranked data, namely at the low and high rank regions, has also been found, for example, in the ranking of cities by size [40]. Of course, the number of COVID-19 preprints and the corresponding rank-size distribution of the repositories are evolving in time. As more data is accumulated, it will be interesting to see whether the above trend continues or whether the points in the low-to-intermediate rank region will get closer to the q-exponential curve, in which case the entire rank-size distribution might be described by a single function.

**Figure 3.**
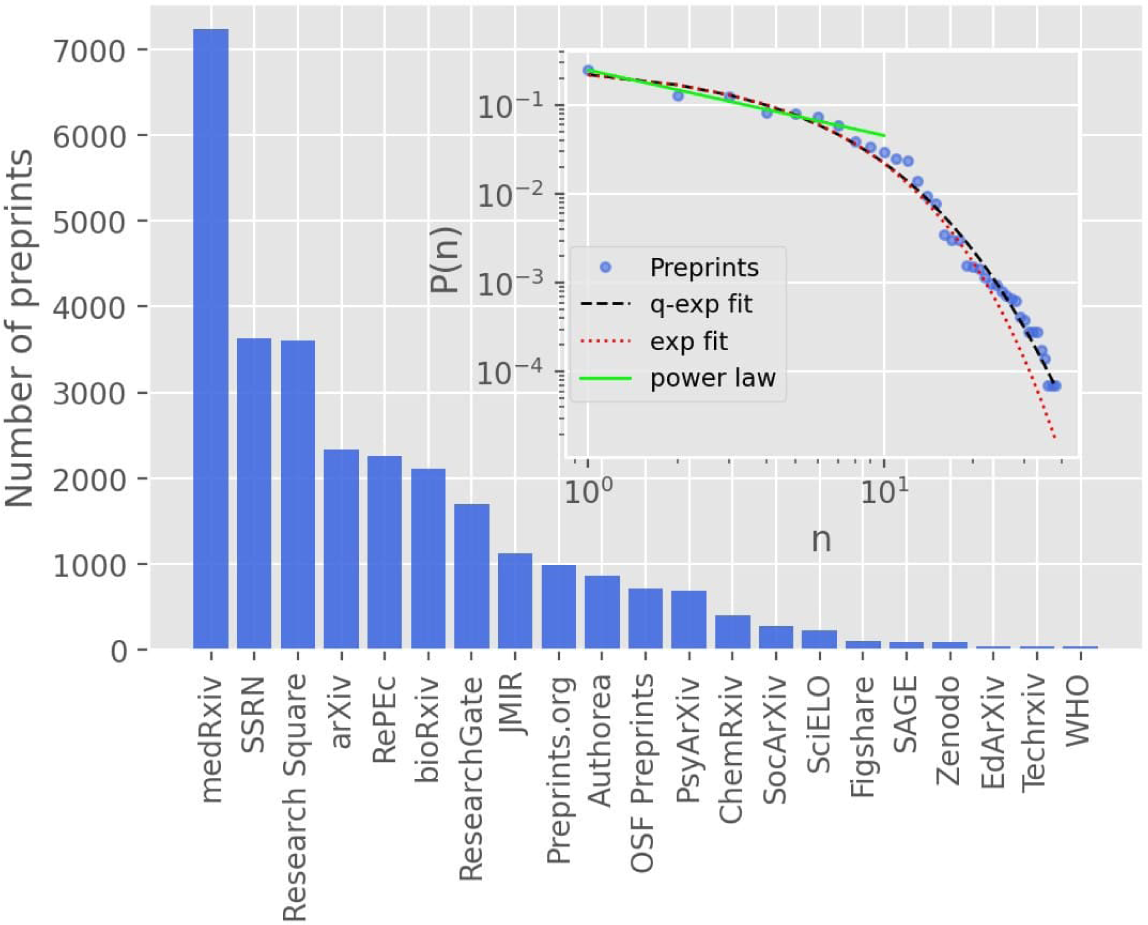
The main plot shows the ranking of the largest preprint repositories by number of COVID-19 related preprints, including all repositories with at least 40 preprints, up to September, 30, 2020. The inset shows the complete rank-frequency distribution in log-log scale, where the red dotted line is an exponential fit, *P* (*n*) ∝ exp(−*αn*), with *α* = 0.25 ± 0.02; the black dashed line is a fit with the *q*-exponential function, *P* (*n*) ∝ exp_*q*_ (−*βn*), where *q* = 1.05 and *β* = 0.27 ± 0.02. The solid green line is a power-law fit, *P* (*n*) ∝ *n*^−*ρ*^, to the first seventh largest repositories, which yields the exponent *ρ* = 0.73 ± 0.06.

## 4 DISCUSSION

We have seen above that the time evolution of the number of COVID-19 related preprints is well described by the generalized Richards growth model. The application of such growth model allows us to infer several interesting aspects of the dynamics underlying the epidemic-like growth of COVID-19 preprints. First, we saw that that early in the epidemic the number of preprints increased in a subexponential manner, as indicated by the value *q* = 0.76 obtained from the GRM fit; see Fig. 1. This means, more concretely, that initially the number of preprints grows polynomially in time according to (6), with an exponent *µ* = 4.2, rather than exponentially fast as was claimed in some early bibliometric studies on the subject [9, 15, 21]. (We recall that pure exponential growth occurs only for *q* = 1.) This subexponential (power law) spreading is also found in many real epidemics [42], including COVID-19 itself [43, 44]. Polynomial epidemic growth is usually attributed to heterogeneous mixing [42, 45], where clustering effects in the underlying propagation network can lead to polynomial spreading [46, 47]. So it is quite likely that such complex dynamics also takes place in the early rapid growth of COVID-19 publications, leading to a subexponential regime.

Another interesting result obtained from Fig. 1 is the fact that the inflection point of the growth profile was reached slightly over four months after the first preprint in our dataset, i.e., *t*_*c*_ = 135 days, corresponding to May 28, 2020, after which the curve has entered a deceleration phase. This deceleration regime can be explained by a combination of factors. First, after a few months of an exceedingly rapid growth in the number of preprints, it becomes naturally more difficult for researchers to obtain novel results at the same pace. Second, it is also possible that after a few months of intensive work on COVID-19, some researchers (especially those whose main areas of expertise are not directly related to epidemics and infectious diseases) may have shifted their focus back to previous research problems or moved on to new ones. Third, starting in late April and early May, repository administrators began to screen more closely COVID-19 preprints against “poor science” [6], and this more stringent vetting processes may have had an impact (however modest) on the rate of accepted submissions. Furthermore, it may also have discouraged authors from submitting manuscripts that they feared would not pass the stricter screening. In other words, enhancing the screening procedures was the equivalent, to some extent, of a ‘mitigation’ intervention in epidemic outbreaks [19]. These factors combined (and possibly others) have lead to a slowing down in the rate of new preprints—an effect that is effectively captured by the saturation term in the GRM, as represented by the term in square brackets in Eq. (1).

We have found that the point of maximum growth rate (i.e., zero acceleration) of the theoretical growth curve happened on May, 28, 2020, as indicated by the yellow vertical line in Fig. 1. After this, the growth dynamics entered a decelerated phase that reached its maximum deceleration on July 23, 2020, as shown by the green vertical line in Fig. 1. After the point of maximum deceleration, the dynamics enters a regime of decreasing deceleration and increasing jerk. From the GRM fit, we have also computed the point of maximum jerk (not shown in Fig. 1) and obtained that it is predicted for September 11, 2020. This implies that after that date the growth curve entered a regime of decreasing deceleration and increasing jerk. One important effect of this increasing jerk is that it contributes to “bend the curve” away from the near-linear growth that is typical of intermediate region around the inflection point *t*_*c*_; see Fig. 1. The growth curve would thenceforth develop a more curved profile and possibly approach a near plateau (assuming that the trend were to continue).

After completion of the first version of the present paper, there happened a slight increase in the number of preprint submissions for the month of October, 2020, in comparison to the previous month. This effect is illustrated in Fig. 4, where we show the monthly numbers of COVID-19 preprints posted on online repositories up to December, 31, 2020, where a secondary small peak in October is clearly visible. This second peak, albeit small, can perhaps be interpreted as a sort of second wave effect, in the sense it might have been a result of the actual second wave of COVID-19 infections that occurred in many countries after August, 2020. In other words, the widespread occurrence of second waves of SARS-CoV-2 infections might have rekindled the interest of the scientific community in the pandemic dynamics, leading to an increase in the rate of COVID-19 preprint submissions. Recently, a generalized logistic model with time-dependent parameters has been shown to describe well the COVID-19 epidemic curves that exhibit secondary waves [22]. It is, therefore, quite possible that a growth model with time dependent parameters may also capture second wave effects in the preprint growth dynamics. An analysis of such a problem is however beyond the scope of the present study.

**Figure 4.**
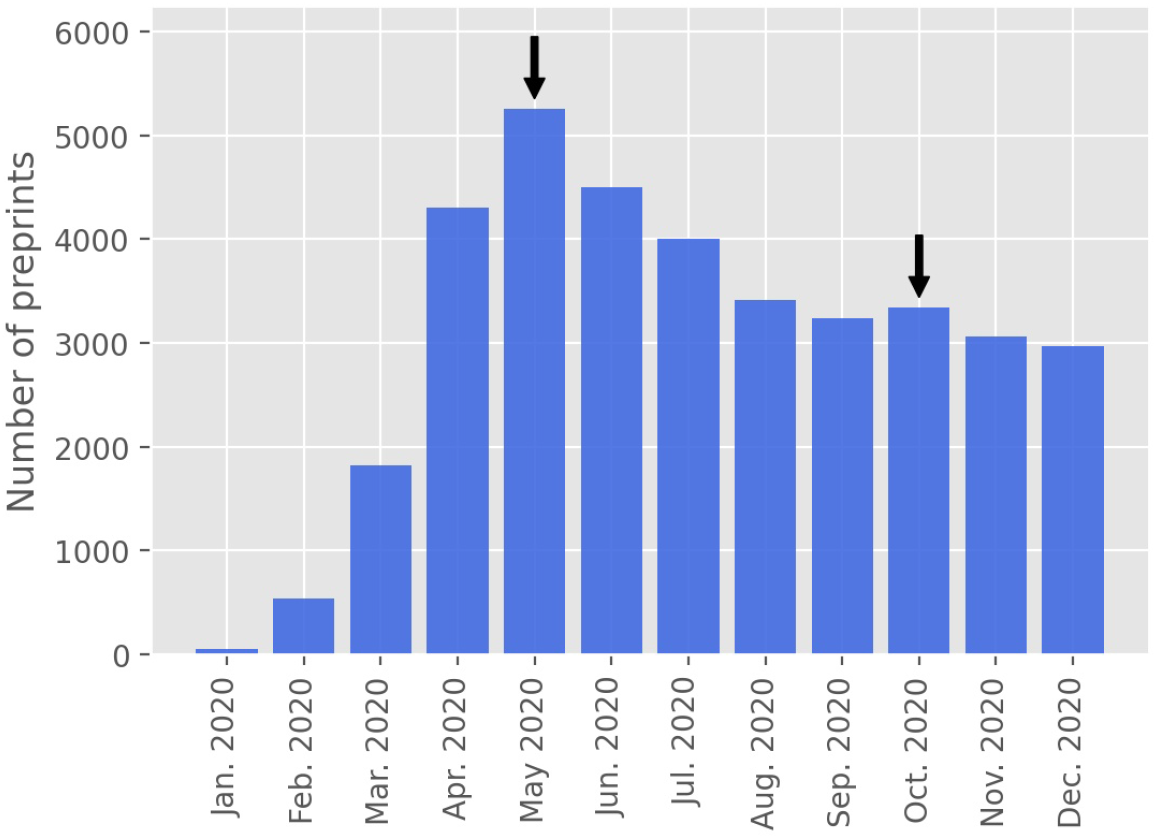
Monthly number of COVID-19 preprints posted on online preprint repositories up to December, 31, 2020. The arrows indicate the main and secondary “peaks” in the graph.

We have also analyzed the size distribution of preprint repositories, as ordered by the number of COVID-19 related manuscripts they host up to September 30, 2020. We have found, in particular, that this distribution is highly peaked at the front-runner (medRxiv)—a property that is typical of the so-called cumulative advantage processes. Such processes often exhibit power-law distributions, and we have indeed verified that the rank-size distribution of preprint servers does appear to have a power-law decay for the first few largest repositories, with a *q*-exponential function describing better the high rank (low size) tail of the distribution. This seems to indicate that a sort of preferential attachment dynamics takes place when authors are considering to which platform to send their manuscripts, as servers that already have more preprints tend to receive more submissions.

It should be emphasized that the problem studied here, namely the growth dynamics of COVID-19 preprints, is a direct manifestation of an underlying complex social process akin to other systems in Sociophysics [26, 36, 37, 38]. It may therefore be possible to develop a microscopic dynamical model, say, at the level of agents, as there is for other social dynamical systems, where the statistical and dynamical trends reported here could be studied in more detail. This is an interesting open problem to be addressed in the future.

In conclusion, it is fair to say that, alongside the public health crisis, the COVID-19 pandemic also triggered a scientific emergency. The scientific community has met this challenge by producing an unprecedented number of scholarly works in a very short period of time, so much so that the phenomenon has been dubbed “an epidemic in an epidemic” [13]. To better understand this scidemic, we have applied a generalized logistic growth model to describe the time evolution of the cumulative number of unrefereed preprints on COVID-19 and SARS-CoV2. Our analysis shows that the quick surge in COVID-19 related preprints can be seen as a sort of contagion process, where existing preprints tend to spur more preprints, and so on, in a cascade-like effect. Eventually this rapid growth is tamed by the system’s own dynamics, as it takes more time and effort for researchers to obtain new results, leading to a deceleration in the growth of the preprint ‘epidemic curve.’ (Other factors, such as closer screening of preprints by the repositories and the return of researchers to pre-epidemic projects, may also contribute to the slowing down in the rate of publication of COVID-19 preprints.) Recently, a possible small second-wave effect on the preprint growth pattern has been observed. This possibility of multiple waves within the COVID-19 scidemic is an interesting question that deserves further examination.

## Data Availability

The datasets analyzed for this study, together with the respective numerical codes, can be found in the website of our research group (http://fisica.ufpr.br/redecovid19/software.html) or can be requested from the authors.

## AUTHOR CONTRIBUTIONS

GLV designed the study. LPC was responsible for collecting and curating the data. LPC, GCDF, and AAB developed the codes. LPC performed the numerical analyses. All authors discussed the results. GLV wrote the first draft. All authors revised and contributed to subsequent drafts. All authors read and approved the final manuscript.

